# B.1.1.7 became the dominant variant in Lebanon

**DOI:** 10.1101/2021.03.17.21253782

**Authors:** Mahmoud Younes, Kassem Hamze, Daniel P. Carter, Karen L. Osman, Richard Vipond, Miles Carroll, Steven T. Pullan, Hassan Nassar, Nada Mohamad, Mohamad Makki, Maysaa Ghadar, Paul Nguewa, Fadi Abdel Sater

**Affiliations:** Research Department, Beirut Cardiac institute, Old airport road, Beirut, Lebanon; Laboratory of Molecular Biology and Cancer Immunology (Covid 19 Unit), Faculty of Sciences I, Lebanese University, Hadath, Beirut, Lebanon; Public Health England, National infection Service, Porton Down, Salisbury, UK; Wellcome Trust Centre for Human Genetics, Nuffield Department of Medicine, Oxford University, Oxford, UK; Bahman Hospital, Beirut, Lebanon; Department of Biochemistry, Oxford University, UK; University of Navarra, ISTUN Instituto de Salud Tropical, Department of Microbiology and Parasitology. Pamplona, Navarra. Spain

**Author notes:** Corresponding Author: Abdel-Sater Fadi: Laboratory of Molecular Biology and Cancer Immunology (Covid 19 Unit), Faculty of Sciences I, Lebanese University, Hadath, Beirut, Lebanon. E-Mail. Co-first authors (Mahmoud Younes, Kassem Hamze).

**Keywords:** B.1.1.7, Emergence, spread, Lebanon

## Abstract

Recently, a new variant of severe acute respiratory syndrome coronavirus 2 (SARS-CoV-2) designated VOC 202012/01 (or B.1.1.7 lineage) has become highly prevalent in several countries, after first being described in the United Kingdom (UK). Its rate of transmission has been estimated to be increased compared to other lineages. In the present study, we show the emergence, dominance and the rapid spread of the B.1.1.7 lineage in Lebanon.

---

Severe acute respiratory syndrome coronavirus 2 (SARS-CoV-2) was, and remains, the most rapidly spreading emerging disease of the year. Recently, a new variant of this virus called SARS-CoV-2 VOC 202012/01 (or B.1.1.7) has been described in the United Kingdom (UK). ^1^ It has become highly prevalent in parts of the UK and is emerging in more than 50 countries. This variant was predicted to potentially be 50 to 70% more rapidly transmissible than other SARS-COV-2 variants circulating in the UK.^2^ The analysis of sequences showed that the new variant had accumulated a large number of mutations that together caused 17 amino acid changes in viral proteins. This variant harbors eight mutations in the spike protein. One of spike mutations is the Δ69-70 deletion in the S1 N-terminal domain (NTD), which has been associated with decreased sensitivity to convalescent plasma.^3^ Other mutations within the spike protein are Δ144, N501Y, A570D, D614G, P681H, T716I, S982A and D1118H. While the others occur in other regions of the genome

## Tracing of B.1.1.7 using RT-PCR

Recently, the European CDC recommended that multi-target RT-PCR assays that included an S gene target affected by the deletions could be used as a signal for the presence of the Δ69-70 mutation to help track these mutant variants.^4^ Multi-target RT-PCR tests using S gene regions impacted by the Δ69-70 mutation are quicker and cheaper than sequencing especially if sequencing capacity is limited. The Applied Biosystems™ TaqPath™ COVID-19 assay allows the detection of this mutation.^5^ In a patient infected with a variant harboring the Δ69-70mutation, there will be an S gene “drop out”. The sequencing of diagnostic PCR amplicon products from S gene dropout samples showed that the S gene target contained the 6-nucleotide deletion (21,765–21,770) encoding the Δ69-70 amino acid deletion within the amplicon. The authors inferred the S gene dropout must be due to a failure of the qPCR probe to bind as a result of the deletion.^6^

## First cases with S gene dropout reported in Lebanon

On December 9, a local patient was diagnosed as COVID −19 positive by PCR using TaqPath 2019-nCoV real-time PCR kit. RDRP and N genes of SARS-COV-2 were detected with a Ct value 18 while S gene was not detected. Two days later, the same profile was detected in two further patients from the same family. The 3 samples were retested using Kylt® SARS-CoV-2 Complete RTU, the results showed detection of IP4 and Spike target genes. These observations showed that the S gene may be affected by the same mutation in these 3 samples. On December 14, the UK health minister first announced that there was a new variant of coronavirus called B.1.1.7. The major mutation in spike gene was the deletion of 6-nucleotides (21,765–21,770) leading to the Δ69-70 deletion. Between 12 and 24 December we detected the S gene dropout in 10 other patients. On December 24, Thermofisher announced that the Applied Biosystems™ TaqPath™ COVID-19 assay, targeting the S gene region impacted by the Δ69-70 mutation, could be used as a signal for the presence of this mutation. Furthermore, we detected an ORF1ab gene dropout in all samples with S gene dropout using primer set (GGTTGATACTAGTTTGTCTGGTTTT and AACGAGTGTCAAGACATTCATAAG) targeting the SGF 3675-3677 deletion in ORF1ab also reported in B.1.1.7.^7^ These findings led us to suggest that our samples with S gene dropout could be the B.1.1.7 variant and that it emerged in Lebanon as early as December 9.

## Rapid Spread of B.1.1.7 in Lebanon

Since December 23rd an increase in the number of patients infected with this new variant has been reported. The first confirmed case infected by the new variant, announced by the Lebanese government, was a passenger arriving from London on December 22. We recorded 498 confirmed cases during the last week of December, among them 52 cases were infected by the new variant. However, in the two first weeks of 2021, the number of cases infected with the new variant increased dramatically. On January 1^st^, 16% of positive cases tested were infected by the new variant. Over a period of 14 days, the number of patients infected with this new variant dramatically increased reaching approximately 60% of positive cases (figure 1). By February 6, the frequency of this variant reached 95% of positive cases. We observed that the rate of positive cases increased from 10% to 25 % in this period. The Lebanese government confirmed 88,392 new cases of COVID-19 and 1,790 related deaths in the country in the 30 days from January 6 to February 6. They imposed a countrywide lockdown to combat the spread of COVID-19. The study was conducted between December 9, 2020 and February 6, 2021. Nasopharyngeal swab samples were collected from 34,978 patients. 6,264 of patients were reported as positive.

**Figure 1.**
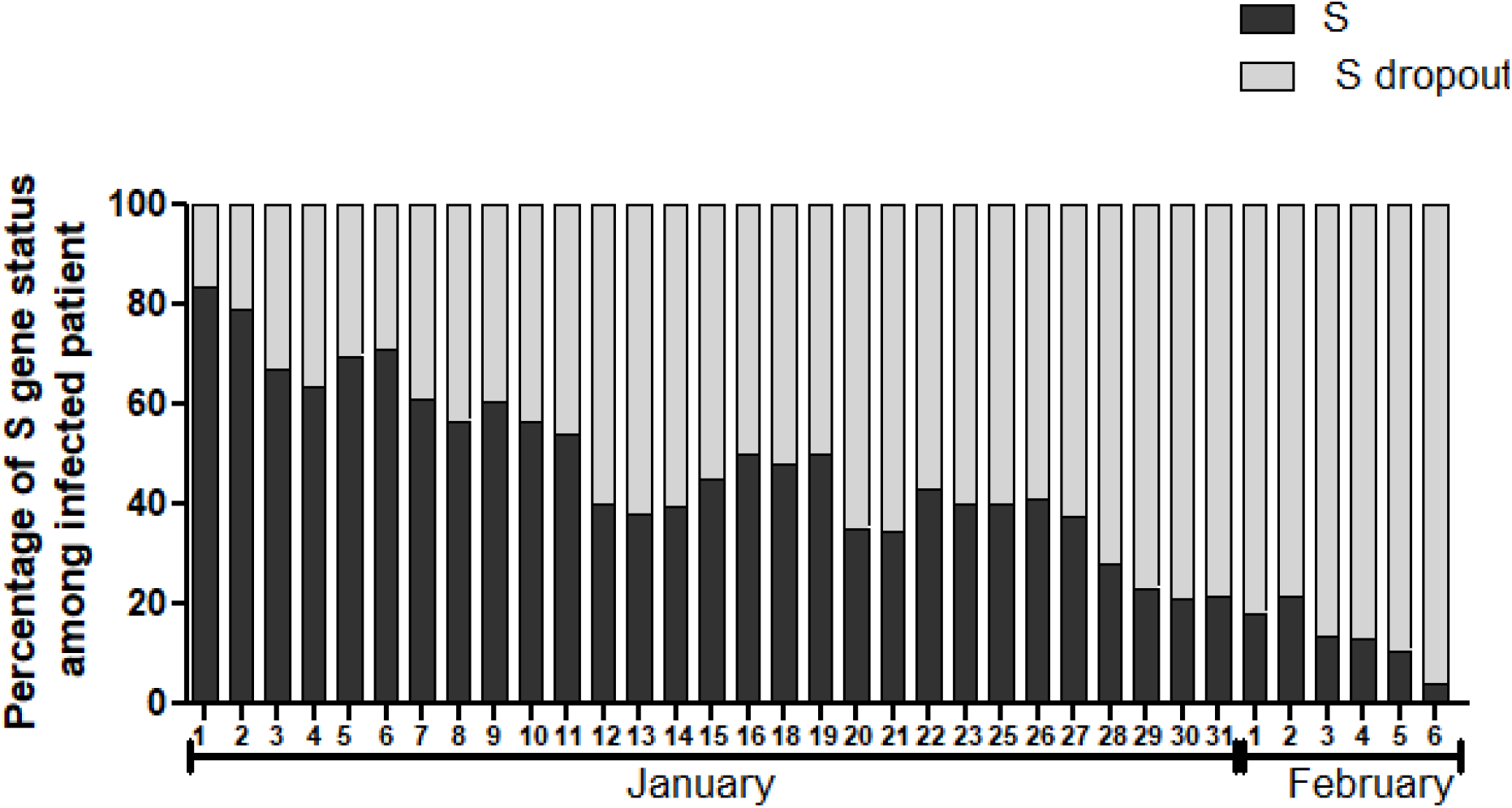
Distribution of infected patients according to the Δ69-70 deletion over a period of 37 days. Of the 19722 patients tested from January 1^st^ to February 6, 4585 were positive, of which 2652 were infected with the new variant. RT-PCR was performed using TaqPath 2019-nCoV real-time PCR kit (Thermo fisher), which targeted the RdRP, N and Spike genes of SARS-CoV-2. All PCR tests were performed using Applied Biosystems™ TaqPath™ COVID-19 assay. S+: spike detection, S dropout: spike not detected;

## Genome sequencing and lineage analysis

To confirm the lineage present in these patient samples, genomic sequencing was performed on nine samples displaying the S-dropout, collected between 9/12/2020 and 8/01/2021. Samples were sequenced using the ARTIC network methodology (https://artic.network/) with the V3 amplicon scheme, V3-LoCost library prep method (Quick 2020) and consensus sequences were generated using the bioinformatics SOP (https://artic.network/ncov-2019/ncov2019-bioinformatics-sop.html) in Nanopolish mode. ^8,9^ Consensus sequences were assigned a lineage using pangolin v2.0 (github.com/cov-lineages/pangolin). Sequences deposited in Genbank under accession MW686007 and MW686013 to MW686019. Sequence 9 deposited in Gisaid under accession number EPI_ISL_1159375. It revealed that eight of the nine were of the B.1.1.7 lineage (Table 1). The remaining sample sequenced belonged to the B.1.258 lineage, which carry the N439K substitution in the Spike protein receptor binding domain (RBD). B.1.258 isolates such as this one, also carrying the ΔH69/ΔV70 deletion, have been reported in several other countries and a proposal has been made to name it B.1.258Δ.^10^

**Table 1.** Genome sequence recovered per sample and Pangolin defined lineage. Presence of the 17 B.1.1.7 lineage SNPS in each consensus sequence. Undetected SNPs in sample 01 and 06 are due to lack of genome coverage in that region.

| Genbank Accession number | Collection date | Percentage of genome recovered | Pangolin defined lineage | B1.1.7 defining SNPS confirmed |
| --- | --- | --- | --- | --- |
| MW686007 | 04/01/2021 | 95.4 | B.1.1.7 | 16/17 |
| MW686013 | 04/01/2021 | 98.1 | B.1.1.7 | 17/17 |
| MW686014 | 08/01/2021 | 98.2 | B.1.1.7 | 17/17 |
| MW686015 | 08/01/2021 | 98.1 | B.1.1.7 | 17/17 |
| MW686016 | 28/12/2020 | 98.1 | B.1.1.7 | 17/17 |
| MW686017 | 28/12/2020 | 97.8 | B.1.1.7 | 16/17 |
| MW686018 | 24/12/2020 | 97.3 | B.1.1.7 | 17/17 |
| MW686019 | 09/12/2020 | 97.9 | B.1.1.7 | 17/17 |
| MW720771 | 12/12/2020 | 86.4 | B.1.258 | N/a |

## Conclusion

The new variant is now the focus of intense debate and analysis, and countries worldwide are scouring their own databases to see if it is circulating within their borders. So far, more than 50 countries have reported the transmission of the B.1.1.7 variant. In the present study, we illustrate the emergence and the fast spreading of a new variant in Lebanon (most probably the B.1.1.7) from December 9.

## Data Availability

data are availabe upon request

